# The COVID-19 pandemic storm in India subsides, but the calm is still far away

**DOI:** 10.1101/2021.06.01.21258143

**Authors:** Igor Nesteruk

## Abstract

In May 2021, the number of new COVID-19 patients in India began to decline, as predicted by the generalized SIR-model (susceptible-infected-removed). The calculations of the final size of this pandemic wave and its duration probably were too pessimistic. New SIR simulations with the use of fresher datasets are necessary in order to update the predictions and to calculate the difference between the registered (laboratory-confirmed) and real number of cases.

## Introduction

At the end of May 2021, the daily number of new laboratory-confirmed COVID-19 cases in India was already half the maximum of 400,000 observed in April, [1]. Such a rapid decline was predicted by the results of preliminary modeling of the epidemic dynamics, published in [2] in early May 2021. Here we will compare the latest data reported from Data Repository by the Center for Systems Science and Engineering (CSSE) at Johns Hopkins University (JHU) [1] with the forecasts based on the generalized SIR-model [2] and will analyze their accuracy.

### Data

We will use the data sets regarding the accumulated numbers of confirmed COVID-19 cases in India (*V*_*j*_) from JHU) [1]. The values *V*_*j*_ and corresponding time moments *t*_*j*_ in May 2021 are shown in Table 1. The values *V*_*j*_ registered in India in March and April 2021 are listed in [2]. The dataset corresponding the period April 10-23, 2021 was used in [2] to calculate the optimal values of the SIR-model parameters. Other *V*_*j*_ values registered in March-May 2021 (in particular, those shown in Table 1) will be used only to check the accuracy of simulations performed in [2].

**Table 1.** Cumulative numbers of laboratory confirmed Covid-19 cases in India *V*_*j*_ according to JHU, [1].

| Day in<br>May<br>2021 | Number<br>of cases,<br>$V_j$ |
| --- | --- |
| 1 | 19557457 |
| 2 | 19925517 |
| 3 | 20282833 |
| 4 | 20664979 |
| 5 | 21077410 |
| 6 | 21491598 |
| 7 | 21892676 |
| 8 | 22296081 |
| 9 | 22662575 |
| 10 | 22992517 |
| 11 | 23340938 |
| 12 | 23703665 |
| 13 | 24046809 |
| 14 | 24372907 |
| 15 | 24684077 |
| 16 | 24965463 |
| 17 | 25228996 |
| 18 | 25496330 |
| 19 | 25772440 |
| 20 | 26031991 |
| 21 | 26289290 |
| 22 | 26530132 |
| 23 | 26752447 |
| 24 | 26948874 |
| 25 | 27157795 |
| 26 | 27369093 |
| 27 | 27555457 |
| 28 | 27729247 |
| 29 | 27894800 |
| 30 | 28047534 |

### Generalized SIR model and parameter identification procedure

In [2] the generalized SIR-model and the exact solution of the set of non-linear differential equations relating the number of susceptible *S*, infectious *I* and removed persons *R* was used (see, e.g., [3, 4]). The exact solution depends on five unknown parameters. The values *V*_*j*_, corresponding to the moments of time *t*_*j*_ from the period April 10-23, 2021 have been used in [2] to find the optimal values of these parameters corresponding to the new epidemic wave in India. The details of the optimization procedure can be found in [5].

## Results and Discussion

The results of SIR simulation performed in [2] are shown in Figure by blue lines: *V(t)=I(t)+R(t)* – solid; dashed one represents the number of infectious persons multiplied by 5, i.e. *I*(*t*)×5; dotted line shows the derivative (*dV* / *dt*)×100 calculated with the use of formula:

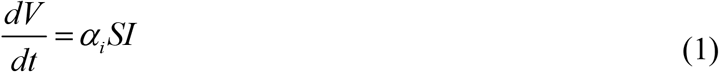

Equation (1) follows from the differential equations of SIR-model (*α*_*i*_ is one of its parameters) and yields an estimation of the real daily number of new cases. Red “Circles” and “stars” correspond to the accumulated numbers of cases registered during the period of time taken for SIR simulations (April 10-23, 2021) and beyond this time period, respectively. It can be seen that these values started to deviate from the theoretical blue solid line in May 2021.

**Figure.**
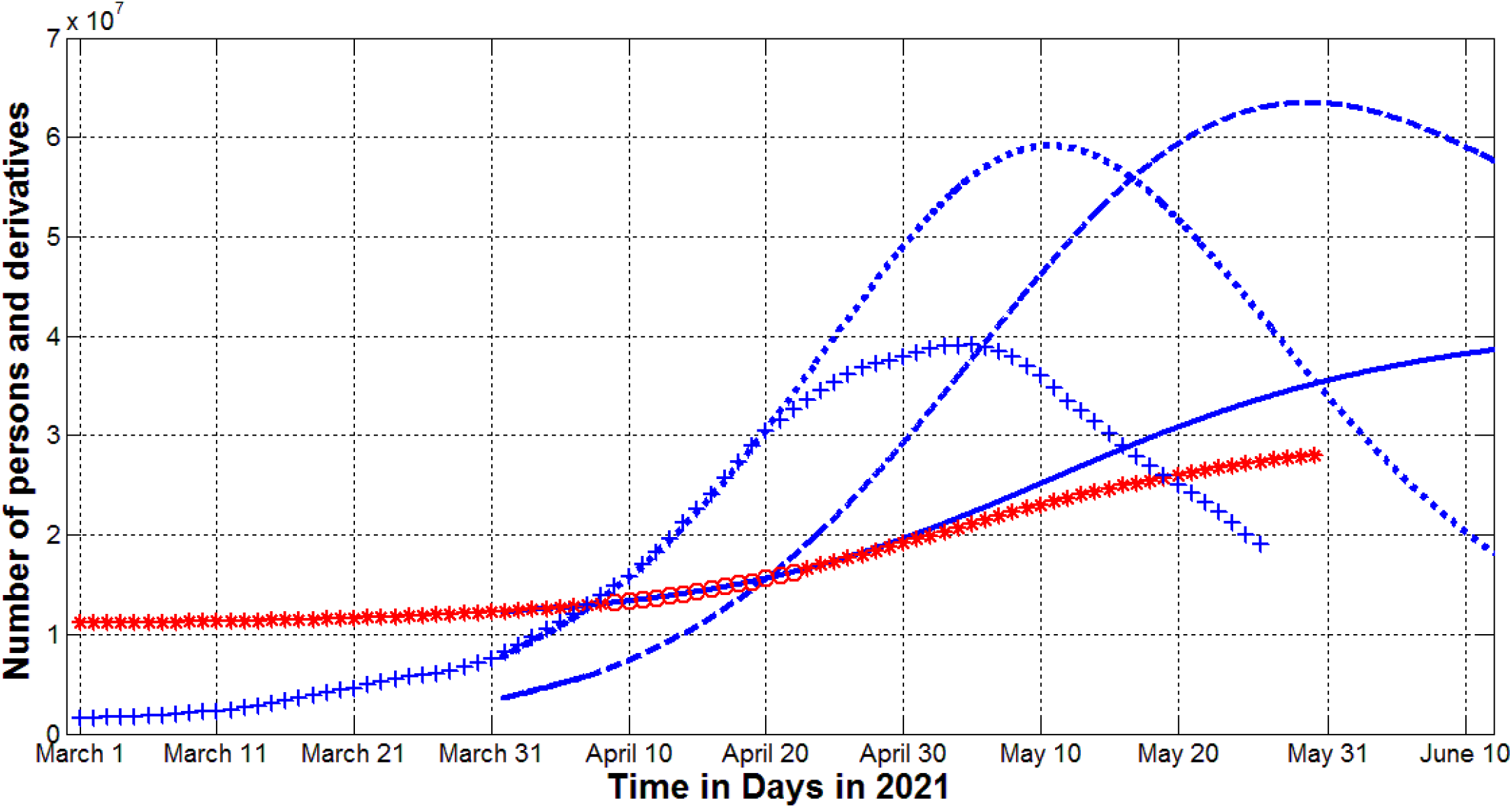
The COVID-19 epidemic dynamics in India. The results of SIR simulations are shown by blue lines. Numbers of victims *V(t)=I(t)+R(t)* – solid; numbers of infected and spreading *I(t)* multiplied by 5 – dashed; derivatives *dV/dt* (eq. (1)) multiplied by 100 – dotted. Red markers show the real number of cases and its derivatives: “circles” correspond to the accumulated numbers of cases taken for calculations; “stars” – number of cases beyond the period taken for calculations (all the values from Table1 and corresponding table in [2]); “crosses” – the first derivative (3) multiplied by 100.

We can also compare the theoretical curve (1) with the average daily number of new cases which can be calculated with the use of smoothing registered *V*_*j*_ values [3, 4]:

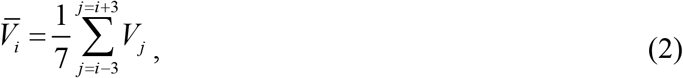

and its derivative:

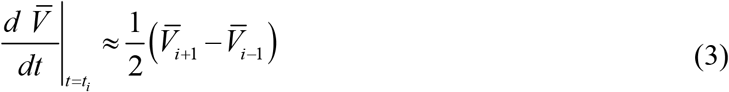

The blue “crosses” represent the results of calculations of the derivative (3) and are in a good agreement with the theoretical dotted line for moments of time before April 22, 2021. After this period we can see a significant discrepancy, but the time moments corresponding to the maximum values of the daily numbers are very close. The maximal value of derivative (3) was achieved approximately one week before the maximum of the theoretical value (1). After two weeks, both the theoretical estimation (1) and the average recorded value (3) decreased approximately twofold.

The revealed discrepancies can be the result the convergence problems mentioned in [2].

Probably new simulation with the use of fresher datasets could fix this problem and provide new more optimistic predictions for the final size and the pandemic duration in India. The second reason for discrepancies may be the large number of unregistered cases observed in other countries [6–9]. Estimates for Ukraine and Qatar made in [10, 11] showed that the real number of cases is probably 4-5 times higher than registered and reflected in official statistics. Similar estimates have to be done for the case of India.

## Data Availability

data is listed in the text

## Acknowledgements

The author is grateful to Oleksii Rodionov for his help in collecting and processing data.

